# Immune molecular profiling of a multiresistant primary prostate cancer with a neuroendocrine-like phenotype

**DOI:** 10.1101/2020.06.01.20115345

**Authors:** Scott G Williams, Han Aw Yeang, Catherine Mitchell, Franco Caramia, David J Byrne, Stephen B Fox, Sue Haupt, Paul J Neeson, Ygal Haupt, Simon P Keam

## Abstract

Understanding the drivers of recurrence in aggressive prostate cancer requires detailed molecular and genomic understanding in order to aid therapeutic interventions. Here we present a case study of a male in 70s with high-grade clinically-localised acinar adenocarcinoma treated with definitive hormone therapy and radiotherapy. The patient progressed rapidly with rising PSA and succumbed without metastasis 52 months after diagnosis. We provide here a description of histological, transcriptional, proteomic, immunological, and genomic features associated with this disease – identifying hallmarks of canonical neuroendocrine PC disease and other novel molecular features.

## Introduction

Prostate cancer (PC) that recurs after primary treatment is a significant health issue. While the initial response rate to conventional therapy is high, most recurrences will progress to become resistant to ongoing salvage therapies, with high rates of lethal progression. More unusually, there are a subset of cancers that are refractory to therapies such as androgen deprivation therapy (ADT) de novo and carry a worse prognosis via rapid progression. Identifying the molecular progression and the genomic features of these tumours will aid in early identification, treatment selection and novel therapeutic development.

Here we present comprehensive histological, molecular and bioinformatic analysis of a case study of a male in 70s initially diagnosed with high grade but clinically localised prostate adenocarcinoma with local disease progression despite combined ADT and radiotherapy (using external beam radiation and brachytherapy), and died from progressive local disease-related complications in the absence of detectable metastases within 5 years of diagnosis.

In this study, we interrogated tissue samples available at baseline as well as multiple stages of the local progression of disease using a combination of histological, immunological, transcriptomic, proteomic, and genomic techniques. This revealed the emergence of an immune-excluded neuroendocrine-like tumour histology harbouring molecular and genomic characteristics of canonical neuroendocrine PC (NEPC). Despite these similarities, we have identified numerous novel molecular features which could improve our understanding of NEPC pathologies.

## Methods

### Tissue collection and processing

Tissue collection methods were varied amongst the four samples and comprised of either transrectal core biopsy (single biopsy 18G; *diagnostic*), transperineal core biopsies (five 16G cores; *RT-* and four 16G cores; *RT+*). The recurrence biopsy was a selected block of FFPE tissue from transurethral resection (TURP; *Recurrence*). Fresh tissue from each patient sample was fixed immediately by immersion in a solution of 10% neutral buffered formaldehyde prior to paraffin-embedding.

### Histology and staining

3 µm sections were collected and used for histopathological analysis including H&E and DAB stains. Staining for neuroendocrine histology used antibodies for synaptophysin (SYN: NCL-L-SYNAP-299, Leica Novocastra), neuron-specific enolase (NSE: Clone BBS/NC/VI-H14, Dako), thyroid transcription factor-1 (TTF-1: Clone SP141, Ventana), chromogranin (CG; Clone DAK-A3, Dako), CD56 (MRQ-42, Cell Marque) and androgen receptor (AR: Clone AR441, Dako). Dual colour IHC was performed on a Ventana Benchmark ULTRA autostainer using CD3 (Clone SP7, Abcam) with OptiView DAB and CD45 (Clone 2B11+DP7/26, Dako) with ultraView Universal AP detection systems

### Immune profiling and cell counting

High resolution scans of dual-color CD3/CD45 stains were uploaded into Halo (Indica Labs) software for densitometry analysis. Whole tissue sections for all four samples were spatially segmented within the software according to histopathological classification of tumor-bearing, tumor-stroma, and mixed tissue zones by a trained pathologist (Figure 2A). Cells were counted within either the tumor stroma or tumor zones according to two cell phenotypes: (i) CD45^+^ CD3^−^, and (ii) CD3^+^, representing non-lymphocytic leucocytes and lymphocytic T cells, respectively. The number of identified cells in each tissue tissue zone was then normalized to the total surface area (Table 1) to account for differences in tissue size.

**Table 1:** Total surface area of tumor and tumor stroma zones in four patient samples.

| Sample | Tissue Zone | Surface Area (mm<sup>2</sup>) |
| --- | --- | --- |
| **Diagnostic** | Tumor stroma | 1.4 |
|  | Tumor | 4.78 |
| **Pre-RT** | Tumor stroma | 2.25 |
|  | Tumor | 8.63 |
| **Post-RT** | Tumor stroma | 5.11 |
|  | Tumor | 9.73 |
| **Recurrence** | Tumor stroma | 7.23 |
|  | Tumor | 22.77 |

**Figure 1:**
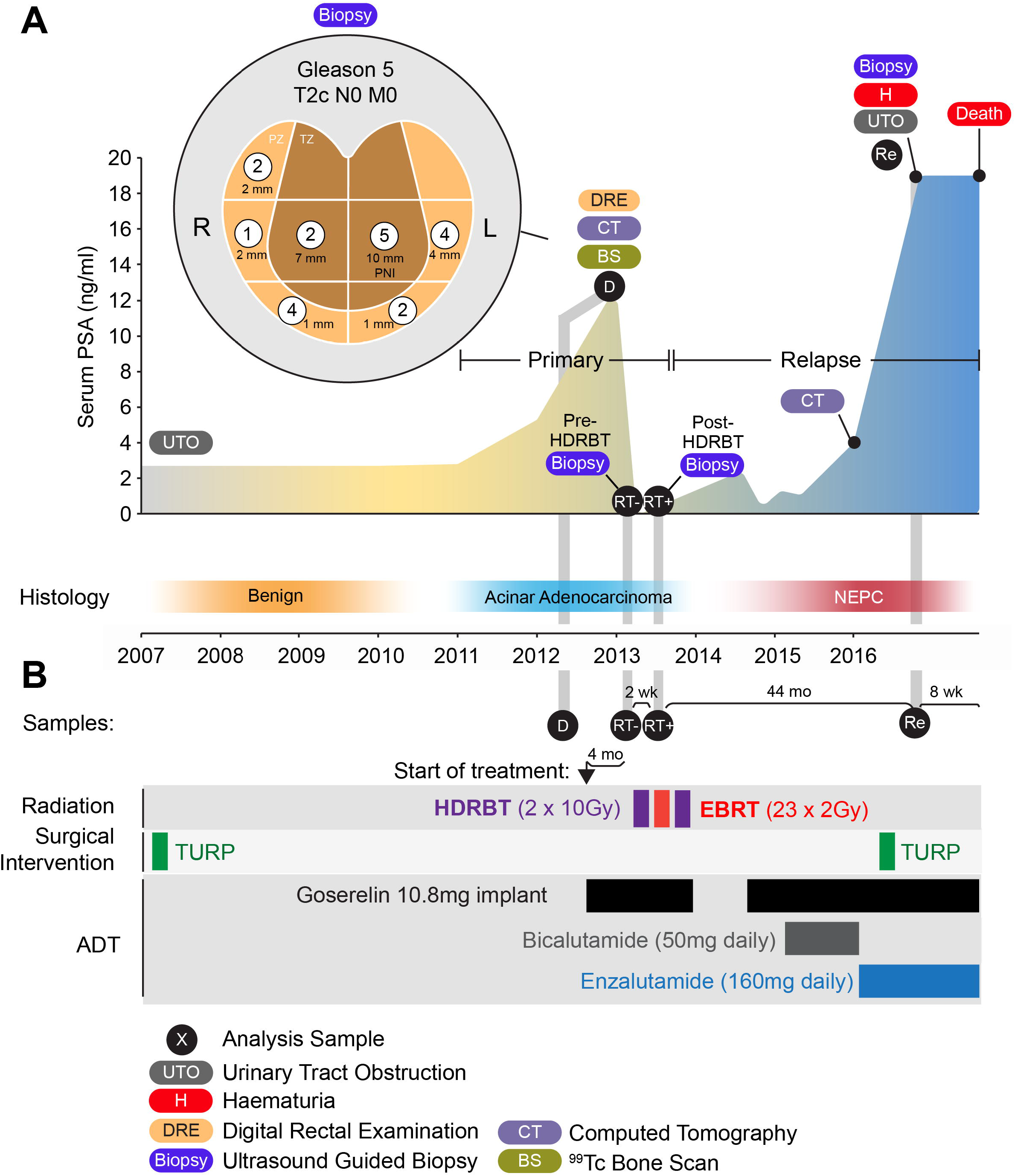
Overview of patient case study. (A) Serum PSA concentration over 10 year course of disease with ultrasound-guided sextant core biopsy results (inset). Numbers indicate dominant Gleason grade in each core with length of tumour involvement indicated in millimetres. Primary disease and relapsed castration resistant disease periods indicated with histopathological classification indicated. (B) Types of duration of treatments performed. HDRBT: high dose-rate brachytherapy, EBRT: external beam radiation therapy, TURP: transurethral resection of prostate. PNI: Perineural invasion

**Figure 2:**
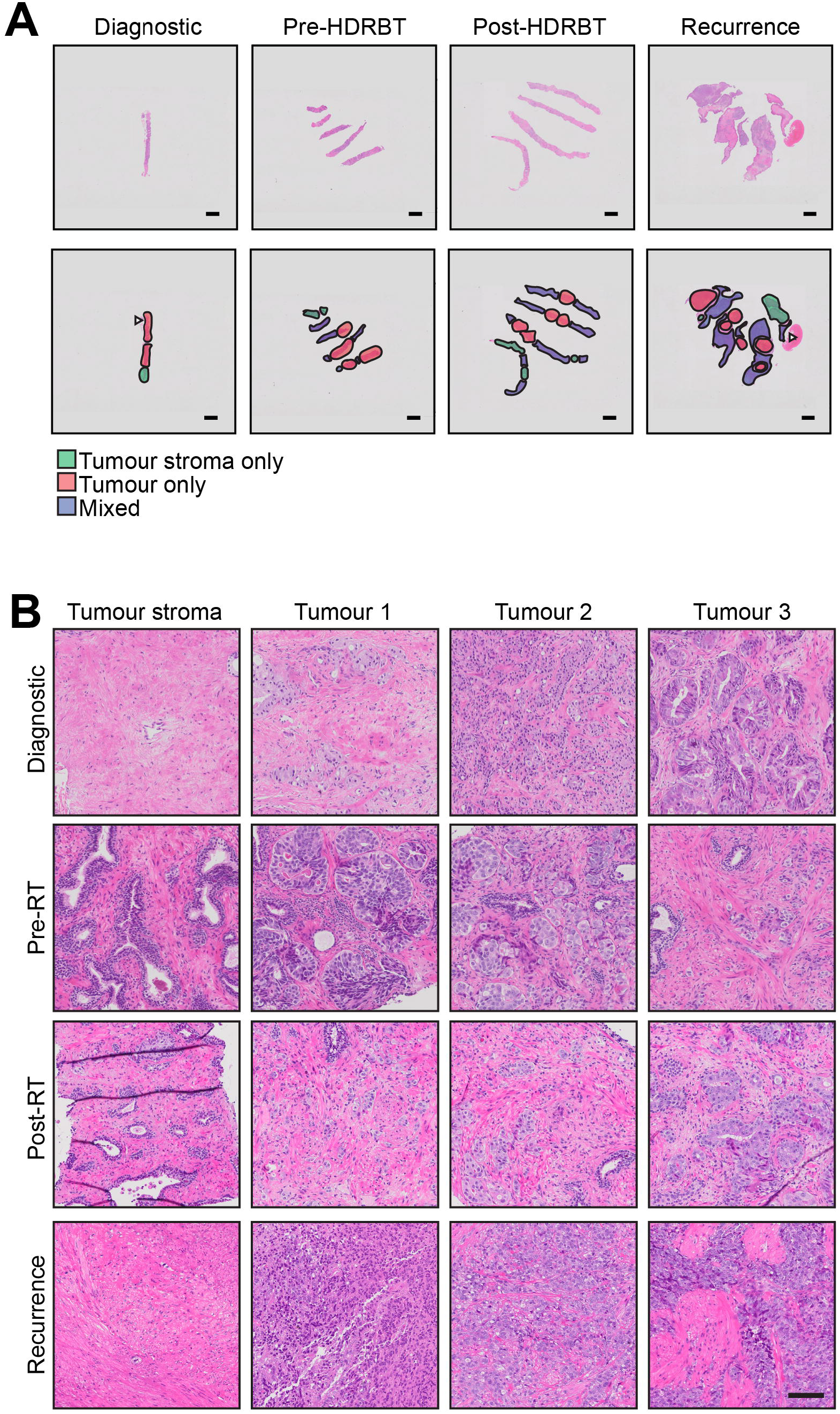
Histopathological classification of tumour and tumour stroma zones identifies initial acinar adenocarcinoma and progression to small cell carcinoma. (A) Overview of H&E stained sections used for histopathological and molecular analysis. Tissue zones comprising (i) tumour, (ii) tumour stroma, and (ii) mixed are indicated. Scale bar indicates 2 mm. (B) High magnification H&E stains of tumour stroma (one zone) and tumour (three zones) at each tissue collection timepoint. Scale bar indicates 100 µm.

### Transcriptomics and proteomics analyses

10 µm sections of formalin-fixed tissue blocks were obtained for downstream RNA and protein extracted for transcriptomic and proteomics analysis, respectively, as per previous studies [1, 2]. This generated normalized mRNA and protein expression levels across the four samples. As input for GSEA analysis of tissue comparisons, normalised data was ranked by magnitude of difference between pairs of samples. For recurrence tissue, all comparisons were made directly to diagnostic tissue (i.e. Recurrence – Diagnostic) in absence of androgen deprivation and radiation therapy.

### Genomic CNA profiling

Total genomic DNA was extracted from 10µm sections of formalin-fixed tissue blocks using QIAamp DNA FFPE tissue kit (Qiagen; Cat: 56404). The total amount of DNA obtained for each biopsy section was 20.7ng (diagnostic), 55ng (RT-), 51.5ng (RT+) and 112ng (recurrence). DNA libraries were prepped for sequencing using NEBNext Ultra II (NEB) kit and sequenced using a NextSeq HO 75PE (Illumina) run. The number of sequences used as input for CNA analysis was 10,815,543 (diagnostic). 6,023,339 (RT-), 8,166,806 (RT+), and 3,983,279 (recurrence).Raw Bam files were analysed for CNA variation using the unaltered R package *QDNAseq* [3]. Parameters for analysis were a binSize of 500 with CNA calls made with “cutoff” method. Called bins were subsequently exported in .igv format for heatmap generation and gene location identification in IGV using genome version hg19.

### Bioinformatic analyses

Fold changes in transcripts/proteins were calculated relative to diagnostic level. Heatmaps were generated using Perseus computational platform using z-score row-normalised log-expression values. Gene set enrichment analysis (GSEA) was performed using Broad GSEA software, and Molecular Signature Database (MSigDB) according to default parameters and established protocols [4]. An FDR of <0.2 was considered significant for GSEA pathway analysis. Output data was visualised using custom *ggplot* R script.

## Results

### Patient description and family history

The case study patient was a male in 70s with pre-existing conditions that were treated with metformin, candesartan and low-dose aspirin. A male relative died of cardiac disease in 50s, and a female relative died from colon cancer in 80s. The patient was a non-smoker with only infrequent alcohol intake. An overview of diagnostic PSA metrics, treatments, pathology and sample acquisition time-points are illustrated in **Figure 1**.

### Diagnosis

Patient had been undertaking PSA screening. In 2007, serum total PSA was 2.70 ng/mL (reference range <4.51 ng/mL) and subsequently was 2.98 ng/mL in 2011. He had developed mild lower urinary tract obstructive symptoms at that stage and proceeded to a transurethral resection of prostate with good symptomatic relief. Histology from that procedure was benign. In 2012, his PSA had risen to 5.29 ng/mL and then 12.81 ng/mL the following year. His urinary function was unchanged but he had a prostate suspicious for malignancy on digital rectal examination; clinical stage T2c. Histology from a transrectal ultrasound-guided biopsy of prostate showed every region contained prostatic acinar adenocarcinoma: Gleason Grade Group (GGG) 1 (right midzone, 2mm of core involved), GGG 2 at right base, left apex and right transition zone (1, 2 and 7 mm respectively), GGG 4 at left midzone and right apex (4 and 1 mm respectively), and GGG 5 at left transition zone (10mm involvement with perineural infiltration noted). A sample used for subsequent molecular analysis was collected at this stage (**Sample 1: *Diagnostic; D***). Staging with CT abdomen and pelvis and ^99m^Tc-bone scan and reported as showing no evidence of metastases. Final clinical stage T2cN0M0.

### Commencement of treatment

Recommended treatment was androgen deprivation therapy (ADT) for a total of 2 years with radical radiotherapy in the form of external beam RT + high-dose-rate brachytherapy (HDRBT). Goserelin 10.8mg implants provided androgen suppression throughout, and radiation therapy was commenced after 4 months of ADT using a sequence of HDRBT, a 2 week break followed by external beam RT (EBRT), another 2 week break and a further HDRBT fraction. HDRBT delivering 10Gy in a single fraction to the prostate with a 3mm margin in all directions apart from 0mm posteriorly using CT planning. EBRT was to the prostate and seminal vesicles with a 10mm margin in all direction except 6mm posteriorly, delivering 46Gy in 23 daily fractions over 4 weeks using CT planning, intensity modulated beams and daily image guidance with 3 intra-prostatic fiducial markers. Two tissue biopsy samples, each consisting of 4–5 16g cores, were collected prior to each HDRBT delivery; HDRBT fraction 1 biopsy representing the effect of 4 months of ADT **(Sample 2: *Pre-HDRBT; RT-*)**, and fraction 2 HDRBT biopsy representing the effect of ADT plus 10Gy HDRBT + 46Gy EBRT **(Sample 3: *Post-HDRBT; RT+*)**.

### Initial response and subsequent rapid development of castration-resistance

Following 3 months of ADT, testosterone was fully suppressed (<0.4 nmol/L) and PSA reduced to 0.131ng/mL. PSA was 0.066 ng/mL 1 month after RT completion. Patient elected not to continue ADT subsequent to RT. Within 15 months of completion of RT, the PSA had risen to 2.41 and goserelin recommenced. Following a PSA nadir of 0.37 ng/mL at 3 months, the PSA rose rapidly. Addition of bicalutamide (50mg daily) resulted in a PSA fall from 1.3 ng/mL to 1.06 ng/ml at 3 months, followed by a rise to 4.0 ng/mL over the next 7 months. Clinical examination suggested ongoing disease in the prostate, and CT chest, abdomen and pelvis showed no evidence of metastases at that point. He was subsequently commenced on enzalutamide (160mg daily) with no impact on PSA level.

### Disease progression and death

Forty-four months since completion of RT, the patient presented with heavy haematuria, urinary obstruction and PSA of 19 ng/mL. This required surgical intervention where a friable bleeding mass obstructing his urethra was resected transurethrally with initial good symptomatic effect. A component of this resection represented the final biopsy sample **(Sample 4: *Recurrence; Re*)**. He represented with further bleeding a month later. Nephrostomy tubes were unable to protect his renal function and he deteriorated clinically and died as a consequence of locally progressive disease 52 months after RT without evidence of metastases.

### Histopathological analysis: Onset of neuroendocrine-like histology at recurrence

Histopathological examination evaluated differences in the series of patient samples and also defined tumour stroma and tumour regions for subsequent analyses **(Figure 2A)**. Acinar adenocarcinoma was identified in the initial diagnostic and subsequent pre- and post-HDRBT tissues, while poorly differentiated adenocarcinoma with areas of neuroendocrine differentiation was identified in the recurrence biopsy (**Figure 2B**). Immunohistochemistry (IHC) for synaptophysin (SYN), neuron-specific enolase (NSE), thyroid transcription factor-1 (TTF-1), chromogranin (CG), CD56 and androgen receptor (AR) in the recurrence biopsy (**Figure 3**) showed weak and/or patchy positive staining for SYN, NSE, TTF-1 and AR. CD56 exhibited non-specific staining and CG was negative. These results support the initial finding of neuroendocrine-like pathology with intact AR expression. Examination of all samples D, RT- and RT+ using synaptophysin IHC (the most consistent marker in sample 4) showed no evidence of NEPC features, suggesting a late emergence of the NEPC-like phenotype(data not shown).

**Figure 3:**
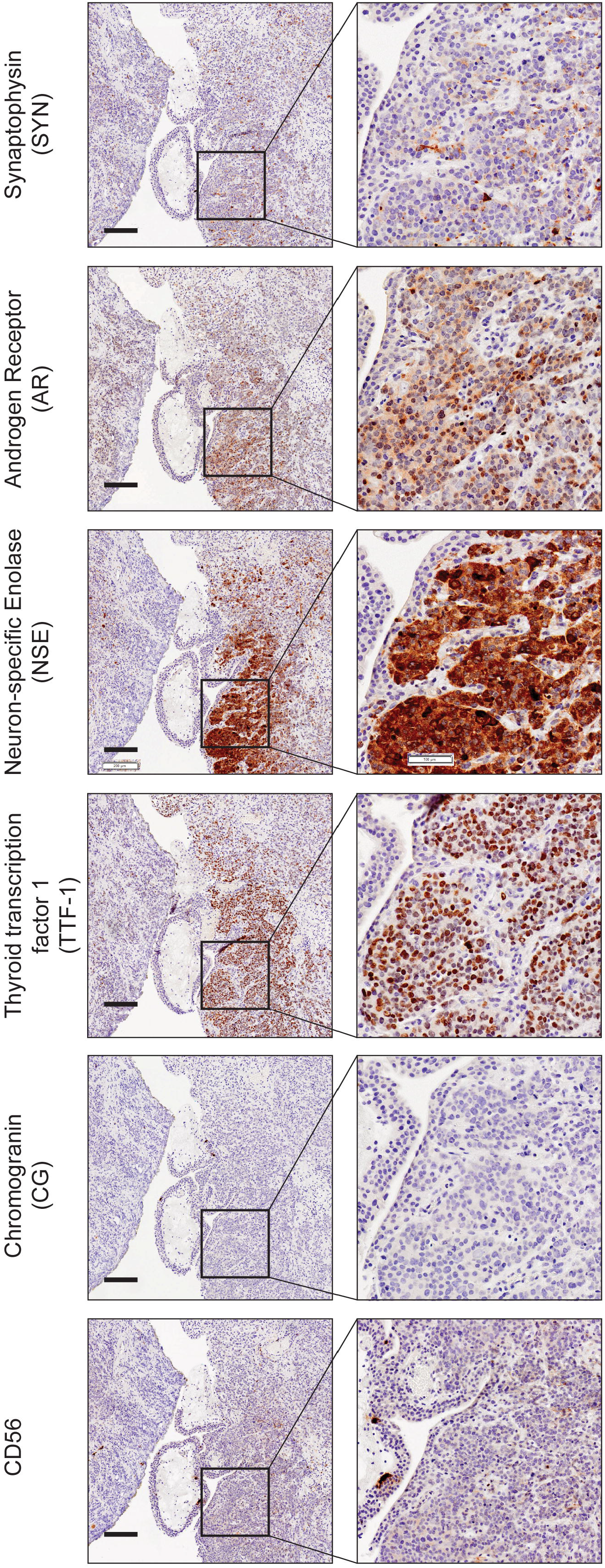
Biomarker staining reveals androgen-intact tumor with NEPC-like marker expression. Serial 3 µm sections from recurrence biopsy sample were stained with antibodies to synaptophysin (SYN), neuron-specific enolase 1 (NSE1), chromogranin (CG), androgen receptor (AR), thyroid transcription factor 1 (TTF1), and CD56. Left panel indicates 4X magnification (scale bar = 200 µm) with box indicating right panel magnification of 15X (scale bar = 100 µm).

### Immune infiltrate is acutely affected by androgen deprivation, radiation and recurrence onset

Studies of the immune context of neuroendocrine cancer are limited to non-existent. We hypothesised that the development of prostatic neuroendocrine-like pathologies could adversely affect the immune context of the tumour and could potentially provide insight into the characteristics of the disease. We therefore performed two-colour dual staining for CD3 (a marker of T cells) and CD45 (a general marker of leucocytes) (**Figure 4A**). We used pathology markup of tumour and tumour stroma zones, as well as unbiased computational calculations of individual cells to generate an overall cell density for both cell types in either the tumour or tumour stroma relative to surface area (**Figure 4B**). Overall, CD3^+^ T cells were far more numerous than other CD45^+^CD3^−^ leukocyte populations. With regards to the effects of acute androgen deprivation (Goserelin for 4 months: D to RT-), we observed that CD3^+^ T cell density was stimulated by up to four-fold (from 116.8 to 414.29 cells/mm^2^) in tumour stroma, but halved (from ∼508.5 to 263.1 cells/mm^2^) in tumour zones. The overall density of CD45+ CD3+ cells was largely unchanged (625.3 to 677.4 cells/mm^2^) from a total tissue perspective. The minor non-T-cell CD45+ CD3-had fewer changes, but did appear to have an increase in density in androgen-treated tissue. Radiation therapy (: RT- to RT+) had the most profound effect on immune cell density, with both populations being heavily reduced post-radiation 2 weeks following the last dose of external beam RT. Finally, we observed that the recurrence sample contained an even lower overall density of immune cells, with around 10-fold less of all cell types when compared to the diagnostic biopsy. Overall, these data suggest that the recurrent PC microenvironment is profoundly immune excluded – regardless of tumour or its surrounding tumour stroma.

**Figure 4:**
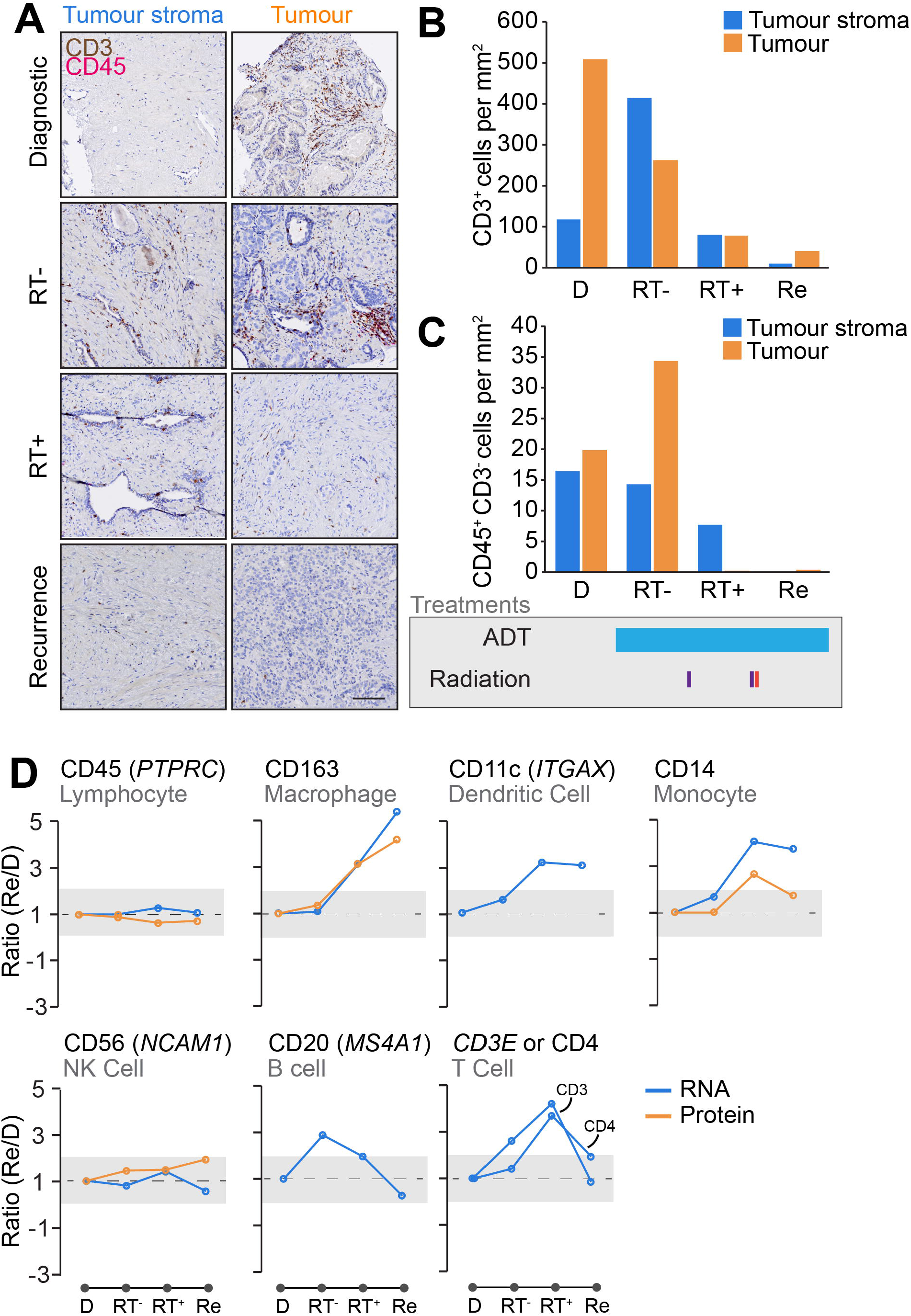
Immuno-transcriptomic profiling reveals steady progression to immune-excluded phenotype. (A) Two-colour (pink and brown) DAB staining of the T cell marker CD3 (brown) and lymphocyte marker CD45 (pink) in four biopsies in tumour stroma and tumour zones. Scale bar indicates 100 µm. Quantification of (B) CD3+ and (C) CD45+ CD3-cell densities (per mm^2^) in each biopsy core tissue zone. Major treatments shown below. (C) Transcriptomic and proteomic analysis of immune cell marker expression changes over course of disease. Shaded areas indicates two-fold change threshold.

Due to limitations on the amount of tissue available for single stain IHC analyses, we used the whole transcriptomic and proteomic data to infer changes in the expression of genes and proteins for markers of major immune cell subsets (e.g. *MS4A1*/CD20 for B cells). The results, shown in Figure 3D, suggested that CD163-expressing macrophages were highly expressed in both post-RT and recurrent tissues. B cells (assessed using *MS4A1* or CD20) responded to early ADT but not to other treatments. Both dendritic cells (*ITGAX*/CD11c) and monocytes (CD14) were stimulated by radiation but plateaued in the recurrent biopsy. Natural killer cells (*NCAM1*/CD56) did not change at any point in disease progression. Assessing T cells (either CD3 or CD4) using this method revealed notable increases after radiation, but drop to diagnostic levels in the recurrent biopsy. These transcriptional changes are not reflected in the previous single-colour stain performed earlier, most likely due to differences in the expression of transcripts and the encoded proteins.

### The transcriptional and proteomic landscape of case study specimens

We performed transcriptomic and proteomic analysis of serial sections taken from all four FFPE biopsies [1, 2] to identify alterations in gene and protein levels (**Figure 5A**). Early changes were consistent with the hormone therapy and radiation treatments. We compared the recurrence (sample 4) to the diagnostic specimen (sample 1) and selected alterations common to both gene and protein analyses – identifying 11 upregulated and 8 downregulated gene/proteins (**Figure 5B**). A description of these molecules and their known links to PC is outlined below.

**Figure 5:**
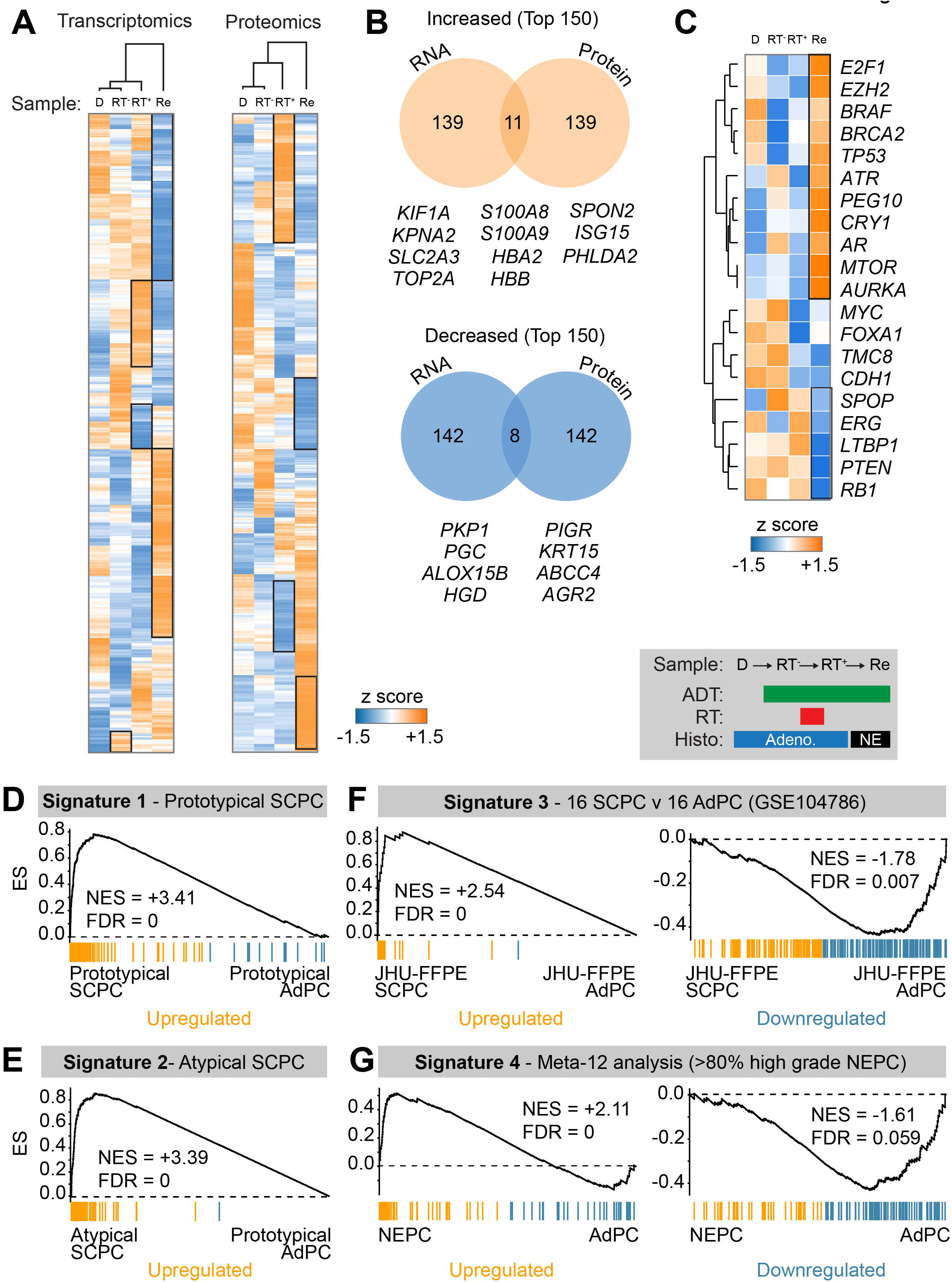
Signature analysis uncovers aggressive SCPC and NEPC genetic profiles in recurrence. (A) Normalised expression heatmaps and hierarchical clustering of transcriptomic and proteomic changes in four biopsies. (B) Venn analysis of overlap in most increased (orange) and decreased (blue) molecules between transcriptomic and proteomic analysis. Common genes are listed below. (C) Normalised transcriptomic expression heatmap of commonly dysregulated genomic lesions in NEPC. (D-G) GSEA comparison transcriptomic changes in recurrence compared to diagnostic biopsy for published prostate cancer gene signatures of (D) prototypical SCPC, (E) atypical SCPC, (F) up- and down-regulated in SCPC versus AdPC and (G) up- and down-regulated in NEPC versus AdPC. SCPC: small cell prostate cancer, AdPC: prostate adenocarcinoma, NEPC: neuroendocrine prostate cancer, FDR: false discovery rate, NES: normalised enrichment score.

#### 1. Upregulated molecules

Within the eleven strongly upregulated molecules, we identified the DNA topoisomerase protein *TOP2A*, which is a known early prognostic biomarker of several high-risk, aggressive and metastatic prostate cancer subtypes [5, 6]. Interestingly, this protein is yet to be linked with neuroendocrine pathologies. We also identified the S100 Calcium Binding Proteins *S100A8* and *S100A9*, which collectively form calprotectin – a faecal biomarker normally associated with intestinal inflammation and previously identified as a faecal marker of radiation-induced proctitis of the prostate [7]. The haemoglobin subunits *HBB* and *HBA2* were also upregulated, likely consistent with the presence of a bleeding mass present at the time of biopsy and noted in the histopathology. Prostatic androgen receptor (AR) targets, including the Kinesin-like protein *KIF1A*, and GLUT3 (encoded by the *SLC2A3* gene) were also observed [8, 9], potentially consistent with the progressively androgen-independent biology of the disease. Karyopherin α2 (*KPNA2*) is a known predictor of biochemical recurrence in prostate cancer treated with radical prostatectomy [10]. Spondin-2, encoded by the *SPON2* gene, has been proposed as a novel serum biomarker of the presence of PC and progression [11, 12]. The interferon stimulated genes *ISG15*, and the PH domain-containing protein *PHLDA2* were also identified. These have no clear link with aggressive PC features in the literature; however the latter is known to be normally expressed in the adult prostate gland (Uniprot: data not shown).

#### 2. Downregulated molecules

Amongst the downregulated proteins identified in recurrence tissue was the anterior gradient protein AGR2 – which has links with various PC pathologies. AGR2 has been shown to be commonly upregulated in primary PC adenocarcinoma, however lower expression levels are observed in metastasis and is highly predictive of biochemical recurrence (BCR) following RP [13]. Multidrug resistance protein 4 MRP4 (*ABCC4*) is an androgen-driven gene that was also lost in the recurrence tissue and is known to be downregulated as PC progresses [14]. Arachidonate 15-lipoxygenase type II (*ALOX15B*), also known as 15-LOX2, is a prostate-specific enzyme also observed to be lost in the recurrent tissue. This enzyme has been shown to be lost in prostate cancer cells relative to normal prostate tissue [15, 16]. Progastricsin, also known as Pepsinogen C, is encoded by the *PGC* gene and also lost in the recurrence sample. Progastricsin is normally found in both normal and malignant prostatic tissue [17]. Its absence could be further evidence of the loss of conventional prostatic biology over the course of the disease. The plakophilin protein PKP1 was also lost and its expression, and it has been already demonstrated to be inversely correlated with Gleason grade in clinical PC adenocarcinomas [18].

### Common putative individual oncogenic drivers of NEPC are altered in recurrence

Candidate analysis did not identify known drivers of lethal PC causing the treatment relapse in this case. As histopathological analysis strongly suggested NEPC biology, we directly tested if common NEPC biomarkers and drivers from the literature (Table 2) were dysregulated in the recurrence. This analysis revealed a strong signature of NEPC-upregulated transcripts (**Figure 5C**) such as *E2F1, AURKA, EZH2, TP53, ATR, PEG10, CRY1, MTOR*. Common genomic losses in NEPC of *RB1, PTEN, LTBP*1, and *ERG* were also reflected in corresponding RNA levels in the recurrent biopsy. These results strongly implicate known genomic and transcriptional drivers of NEPC in mediating the treatment-refractory disease course in this case.

**Table 2:** Known genomic drivers of NEPC.

| *Gene* | *Product* | *Reference* |
| --- | --- | --- |
| *RB1* | RB Transcriptional Corepressor 1 | [19-21] |
| *PTEN* | Phosphatase and tensin homolog | [20] |
| *CHD1* | Chromodomain Helicase DNA Binding Protein 1 | [19] |
| *TP53* | Tumor Protein P53 | [19, 21] |
| *BRCA2* | BRCA2 DNA Repair Associated | [19] |
| *AR* | Androgen Receptor | [19, 22] |
| *SPOP* | Speckle Type BTB/POZ Protein | [19] |
| *TMC8* | Transmembrane Channel Like 8 | [19] |
| *FOXA1* | Forkhead Box A1 | [19] |
| *LTBP1* | Latent Transforming Growth Factor Beta Binding Protein 1 | [19] |
| *CRY1* | Cryptochrome Circadian Regulator 1 | [19] |
| *BRAF* | B-Raf Proto-Oncogene, Serine/Threonine Kinase | [19] |
| *MYC* | MYC Proto-Oncogene, BHLH Transcription Factor | [22, 23] |
| *ERG* | ETS Transcription Factor ERG | [24] |
| *AURKA* | Aurora Kinase A | [22] |
| *E2F1* | E2F Transcription Factor 1 | [25] |
| *ATR* | ATR Serine/Threonine Kinase | [26] |
| *MTOR* | Mechanistic Target Of Rapamycin Kinase | [27] |
| *EZH2* | Enhancer Of Zeste 2 Polycomb Repressive Complex 2 Subunit | [22, 28] |
| *PEG10* | Paternally Expressed 10 | [29] |

### Recurrent disease shares known transcriptional characteristics of prototypical and atypical small cell prostate carcinoma, and NEPC

We next performed ontology enrichment analysis using genesets that are over- or under-expressed in prototypical adenocarcinoma (AdPC), small cell prostatic carcinoma (SCPC), and neuroendocrine prostate cancer (NEPC) published by Tsai *et al*. [30]. For this purpose, we directly compared RNAseq data from the recurrence biopsy with the diagnostic specimen. This enabled a clear comparison of tumour profile without contamination from treatment variables. The results revealed a highly significant signature for both prototypical (**Figure 5D**) and atypical SCPC (**Figure 5E**) when compared to prototypical AdPC. In addition, contrast with another FFPE dataset compared SCPC with AdPC revealed strong signatures for SCPC as a whole (**Figure 5F**). Finally, comparison with a curated genelist specific to NEPC (Meta-12 analysis – present in > 80% of NEPC) again strongly supported an NEPC transcriptional profile (**Figure 5G**). Taken together these data confirm suspicion of NEPC biology. However, due to the known intratumoral heterogeneity within NEPC, and the absence of appropriate comparative datasets, it is difficult to further resolve any of the specific NEPC subtypes any further.

### Aggressive recurrent disease is driven primarily by conventional molecular pathways

Due to the particularly aggressive nature of the recurrent tumour, we sought to identify pathway over-activity (or inhibition) that was likely to be driving the disease. We therefore focused on comparing our data to canonical pathways using Gene Set Enrichment Analysis (GSEA). We interrogated both the more expansive Pathway Interaction Database (PID), and the highly curated MSigDB Hallmark (Hallmark) collections using both the transcriptional and proteomic datasets previously generated. Of particular interest were pathways regulated by known drivers (c.f. **Figure 5C**), and those significant in *both* transcriptomic and proteomic analyses. It should be noted that the proteomic analysis (5,566 proteins) was restricted due to a lower number of identified molecules compared to transcriptomics (12,661 genes).

From the PID analysis, we confirmed that the AURORA Kinase A and B, MYC, ATR/ATM, RB1, E2F and p53 pathways were all upregulated at the transcriptional level (**Figure 6A**). Amongst these, we also identified E2F and ATR upregulated at the proteomic level. Interestingly, we additionally identified the c-Myb transcriptional network upregulated at both the transcriptional and proteomic level. This proto-oncogene pathway has not yet been strongly linked with NEPC, but does have known hyperactivity in the castration-resistant PC adenocarcinoma cell C4–2 [31]. To provide additional evidence for the most important up- and down-regulated pathways, we performed a similar analysis using the Hallmark datasets. The results, shown in **Figure 6B**, identified that E2F, MYC and p53 pathways, MTORC1, TNF-alpha, G2M checkpoint and complement pathways are robustly upregulated. Taken together, these two analyses suggested that Aurora kinase, E2F, MYC and p53 activity, are the putative drivers of the recurrent NEPC pathology.

**Figure 6:**
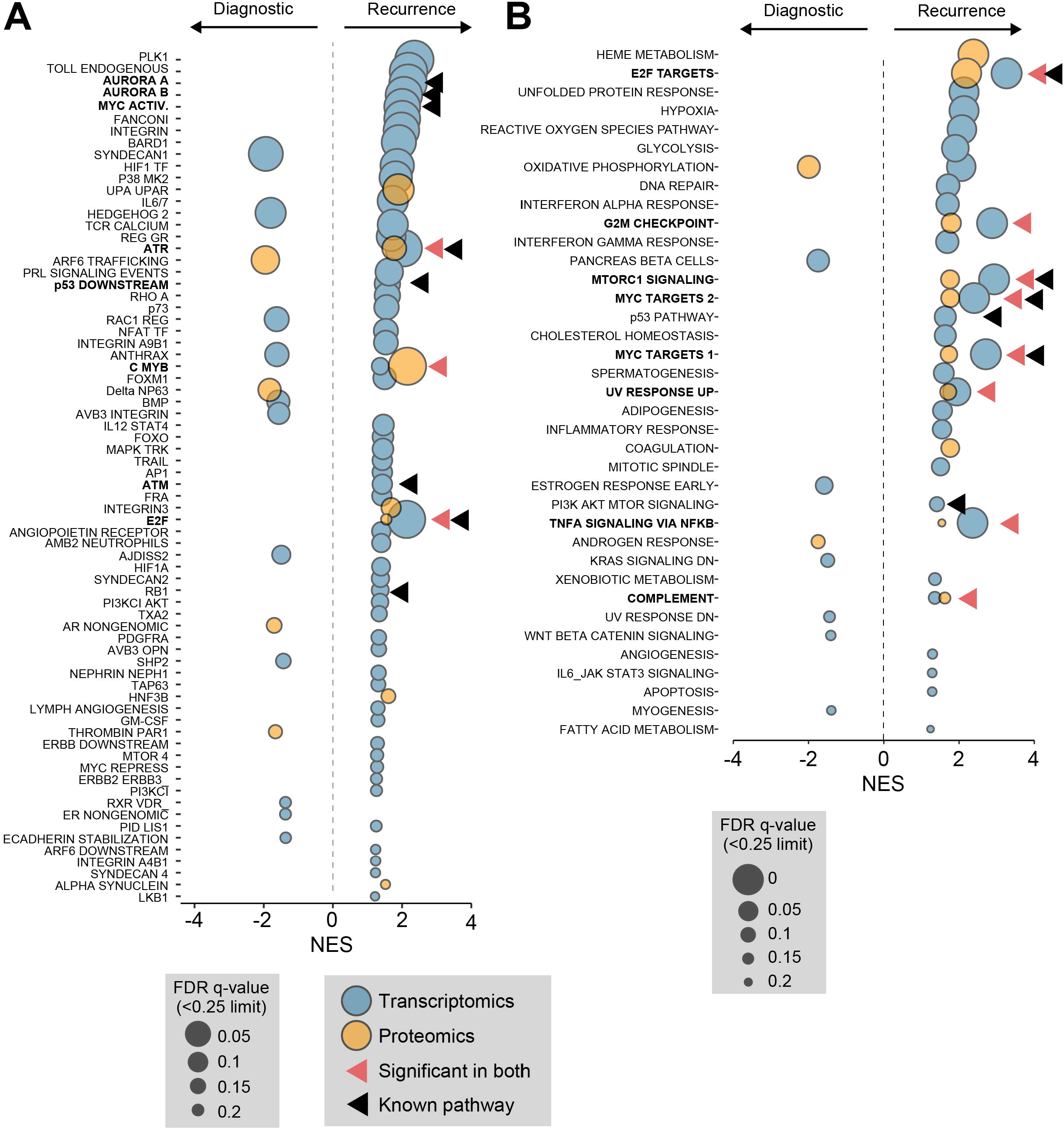
Gene set enrichment analysis of transcriptomics and proteomics revealed canonical NEPC pathway dysregulation in recurrent tumor tissue. Bubblechart of significant enrichment hits of pathways from (A) Pathway Interaction Database (PID) and (B) MSigDB Hallmark database. Red arrows indicating significant hits in both transcriptomics and proteomics. Black arrows indicate pathways regulated by known NEPC drivers. NES: normalized enrichment score, FDR: false discovery rate.

### Whole genome sequencing reveals pervasive chromosomal alterations in recurrent tissue

We identified a total of 18 single-copy copy number alterations (CNAs) in the recurrence which did not appear in any of the preceding samples (**Figure 7A**). These changes included three whole chromosomal arm gains (4p, 5p and 8q) and five losses (4q, 8p, 13q, 17p and 18p). Three smaller amplifications and seven losses were observed elsewhere in the genome (Figure 6; bottom panel). Due to the lower quality and quantity of FFPE material in preceding samples, the analysis could not automatically identify chromosomal changes in these samples. However, several of the recurrence sample CNAs could be observed in the diagnostic biopsy via processing (Figure B). This included the 8p loss / 8q gain, the 12q14.2–24.32 gain, and the 16q12.1–24.2 loss (marked in arrows). Interestingly, these were not as prominent in the post-HDRBT biopsy – potentially due to the elimination of many of the tumours due to irradiation. Both 8p loss and 8q amplification are highly common genomic alterations in all stages of PC – with frequencies of ∼34% and ∼29%, respectively [32].

**Figure 7:**
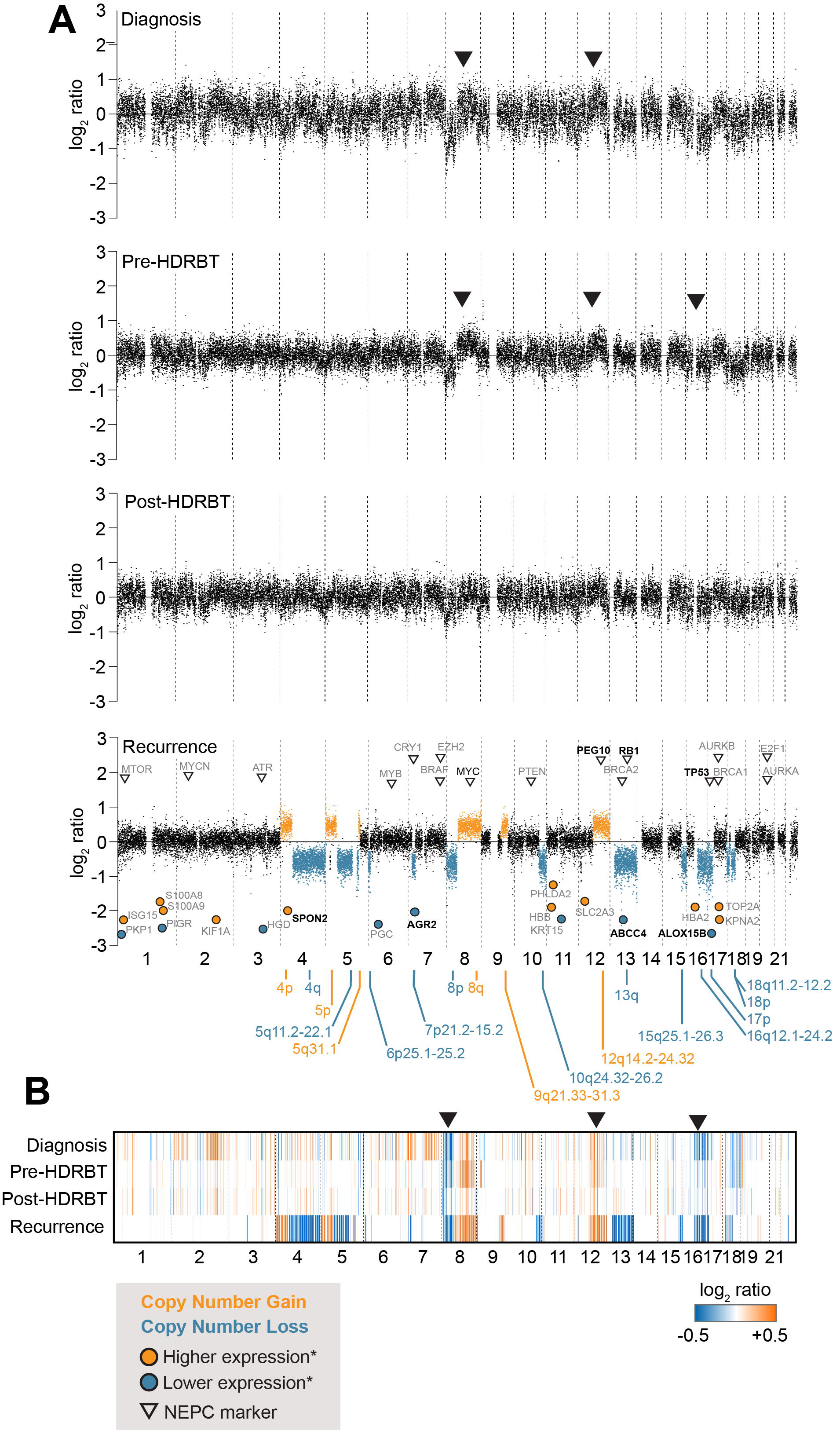
Genomic copy number analysis (CNA) reveals both canonical and novel genomic alterations in emergent NEPC-like tumor. (A) Copy number scatterplot profile and (B) heatmap analysis of low coverage genomic sequencing and copy number analysis of four tissues in biopsy series. Recurrence panel overlayed with location of genes with expression changes from previous analyses, and known NEPC markers (black arrows).

We also observed that several of the highly significant transcript and proteomic changes, as well as known NEPC markers, corresponded to these genomic changes in the recurrent tissue. For molecules already linked with NEPC, these included *PEG10, RB1* and *TP53*. Whilst *RB1* exhibited the same directionality change as expected, the others were not as expected. For example, *TP53* is located on the partially lost chromosomal arm 17p, yet exhibits transcriptional amplification. As previously mentioned, this chromosomal loss also appears to be an early event and visible in the diagnostic tissue. The discrepancy could be due to the subsequent mutation and stabilisation of *TP53*, which has been observed previously in small cell carcinomas of the prostate [33]. A similar observed was also made for *PEG10*, a molecule strongly associated with treatment-induced NEPC [34] and also incidentally requires *TP53* and *RB1* dysregulation [29]. As many of these putative NEPC markers are derived from genomics, rather than transcriptomics, it may be more informative to consider genomic CNAs than transcriptional change.

### Signatures of treatment delivery indicate a conventional response to radiation and ADT

A conceivable source of treatment relapse is inefficacy of the two treatments used. Specifically, suboptimal treatment delivery or effect that leads to progression and the development of castrate resistance. To address this question, we used GSEA to compare the transcriptional and proteomic data at different treatment stages with existing genesets associated with canonical biological responses to (i) ionizing radiation in PC, and (ii) androgen deprivation. For radiation therapy, we drew upon an existing 300-gene (150 up and 150 down) signatures of both the transcriptional [1] (**Figure 8A**) and the proteomic [2] radiation response in PC (**Figure 8B**). In addition, we also compared with our pan-tissue dosimetric transcriptional signature and a corresponding non-dosimetric control signature (**Figure 8C**) [35]. Taken together, the three analyses suggested that the case study exhibited a molecular response to therapeutic radiation in the prostate was consistent with an appropriate radiation treatment response.

**Figure 8:**
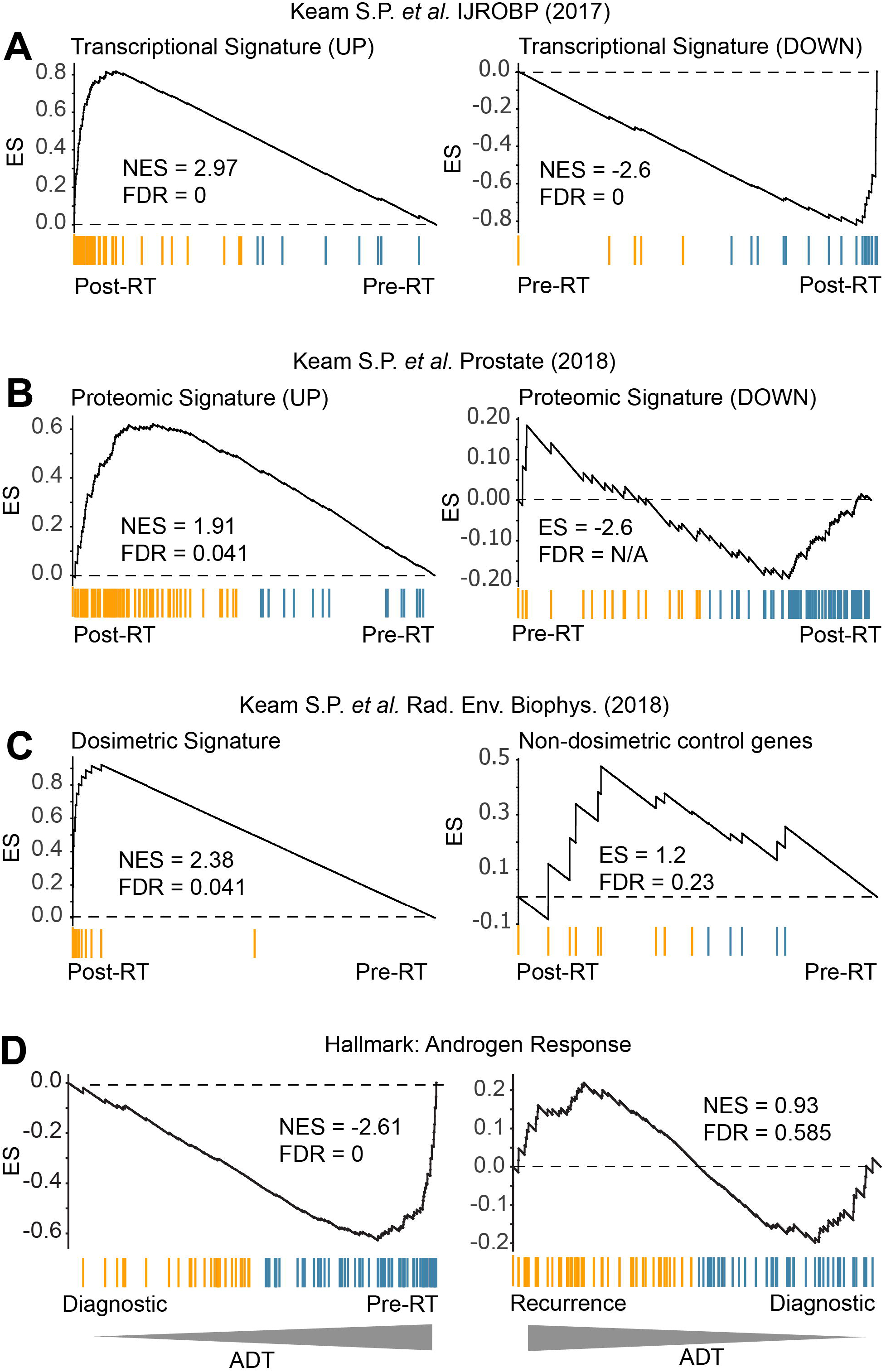
GSEA analysis of biopsy pairs reveals conventional treatment responses. Gene set enrichment analysis was performed on biopsy pairs in order to identify the patterns associated with known radiation and androgen-associated gene and protein expression profiles. (A) Transcriptional and (B) proteomic response to HDRBT from Keam *et al*. 2017 and Keam *et al*. 2018a. (C) Dosimetric geneset response from Keam *et al*. 2018b. (D) MSigDb Hallmark Androgen Response signature. NES: normalised enrichment score. FDR: false discovery rate.

The patient was also treated with androgen-deprivation in the form of goserelin prior to RT, and subsequent to radiation with bicalutamide, and then enzalutamide as CRPC emerged. To test if the initial ADT was effective, we contrasted the pre-RT sample (after 4 months of goserelin) with the baseline diagnostic biopsy (no ADT) at the transcriptional level using known androgen-regulated genes as a proxy for the absence of androgen signalling in the tissue – a consequence of androgen withdrawal. Androgen signalling was inferred using the *Androgen Response Hallmark* gene dataset and analysed using GSEA analysis (**Figure 8D; left panel**). The results revealed the strong downregulation of the androgen pathway in the initial goserelin-treated sample – consistent with the use of goserelin and a biochemical PSA drop from 12.81ng/mL to 0.131ng/mL. This response was in stark contrast to that seen in the recurrence versus diagnostic analysis (**Figure 8D; right panel**) where androgen signalling was no longer significantly altered – reinforcing the androgen-indifferent progression of the relapsed disease despite the administration of potent combination androgen suppression. Together, these results confirm that initial treatments impacted as expected at the molecular level, and do not suggest any atypical therapeutic responses were responsible for the development of aggressive disease.

## Discussion

In this study, we have presented an in-depth molecular case study of a fatal treatment-emergent NEPC-like prostate cancer. Here, initial diagnosis of T2c Gleason grade group 5 disease was subsequently treated with ADT and high-dose-rate brachytherapy/external beam RT. However, a poorly differentiated adenocarcinoma with NEPC differentiation became apparent following therapy and was resistant to increasingly aggressive ADT intervention (goserelin plus bicalutamide or enzalutamide). Our study has identified the cancer possessed some of the classical molecular, histological and genomic hallmarks of small cell PC and NEPC (e.g. Rb loss, AURKA gain etc.), however it did not exhibit many of the key clinical features such as low PSA rise and metastasis [36]. Conversely, we identified many novel molecules that are specifically expressed in the recurrent tissue which have strong links to more conventional PC pathologies and progression (e.g. *ABCC4)*. Many of these new molecules have limited known functional impact on PC, and have instead been defined as novel prognostic or tumor markers. Otherwise, we have identified the c-Myb pathway as being of strong interest for this type of aggressive PC biology – with hyperactivation identified using pathway analysis. However, the literature describing the implication of this pathway in PC is limited to in vitro studies [31]. Further exploration of this pathway in NEPC-like pathologies is warranted. We have also described the lack of an active immunological landscape within this this type of tumor, suggesting that it would not be amenable to any current form of immune-based therapy. Finally, we showed that the molecular response of the tumor to initial radiation- and androgen-based treatments was consistent with previously studied patients; advocating that treatment efficacy was not a factor in the emergence of aggressive disease. The cataloguing of these molecules features in this interesting disease case will potentially provide valuable insight into other rare PC pathologies and improve treatment success.

## Data Availability

Data are available upon reasonable request.

## Declarations

### Ethics approval and consent to participate

The participant provided consent covering tissue research as part of a prospective tissue collection study for prostate radiobiology research approved by the Human Research Ethics Committee at the Peter MacCallum Cancer Centre (PMCC; HREC approvals 10/68, 13/167, 19/32).

### Availability of data and material

The datasets used and/or analysed during the current study are available from the correspond ing author on reasonable request.

